# Association between reactogenicity and SARS-CoV-2 antibodies after the second dose of the BNT162b2 COVID-19 vaccine

**DOI:** 10.1101/2021.07.19.21260744

**Authors:** Shohei Yamamoto, Ami Fukunaga, Akihito Tanaka, Junko S. Takeuchi, Yosuke Inoue, Moto Kimura, Kenji Maeda, Gohzoh Ueda, Tetsuya Mizoue, Mugen Ujiie, Wataru Sugiura, Norio Ohmagari

**Author notes:** **Address for correspondence:** Tetsuya Mizoue, Department of Epidemiology and Prevention, National Center for Global Health and Medicine, 1-21-1, Toyama, Shinjuku-ku, Tokyo, 162-8655, Japan. Equal contribution as first author (Shohei Yamamoto, Ami Fukunaga).

## Abstract

High vaccine reactogenicities may reflect stronger immune responses, but the epidemiological evidence for coronavirus disease 2019 (COVID-19) vaccines is sparse and inconsistent. We observed that a fever of ≥38□ after two doses of the BNT162b2 vaccine was associated with higher severe acute respiratory syndrome coronavirus 2 (SARS-CoV-2) spike IgG titers.

## Introduction

A SARS-CoV-2 vaccine based on mRNA technology, developed by Pfizer and Moderna, has shown 94–95 % efficacy in preventing COVID-19 [1, 2]; the second dose of this mRNA vaccine substantially increases antibody titers of the SARS-CoV-2 IgG spike proteins,[3] while accompanying much greater side effects than that of the first dose [1, 2]. The hypothesized mechanism underlying this phenomenon is that the type I interferons and multiple pro-inflammatory cytokines and chemokines produced by the booster vaccine, which induce injection-site and systemic inflammation (side effects), promote an antibody-producing immune response [4]. The vaccine reactogenicity may thus correlate with the immune response.

However, epidemiological evidence on the relationship between reactogenicity and immunogenicity of COVID-19 vaccines is limited and inconsistent [5-7]. A Japanese study among healthcare workers (HCWs) observed higher levels of IgG spike antibodies after the second dose of the BNT162b2 vaccine among those who experienced systemic symptoms [7]. In contrast, a Korean study of HCWs showed no difference in IgG spike titers according to local and systemic reactogenicity grades after the first and second dose of the AZD1222 or BNT162b2 vaccines [5]. However, no studies have reported the immunogenicity in relation to post-vaccination fever, which is an objective measure of systemic reactogenicity as well as an indicator of immunity activation [8, 9]. Here, we report SARS-CoV-2 IgG spike antibodies in relation to reactogenicity following SARS-CoV-2 vaccination, with particular attention to the onset of post-vaccination fever.

## Methods

We performed a repeated survey after the start of the vaccination program in March 2021 in a cohort of staff members of the National Center for Global Health and Medicine (NCGM), a national medical institution in Japan. Details of the study design are explained in the Supplementary appendix 1.

We recruited 100 staff members who were not taking immunosuppressive medication. Out of the 100 participants, 12 members were excluded who lacked data on symptoms and fever after vaccination, and 88 members were involved for the analyses. The participants were vaccinated with the mRNA-based SARS-CoV-2 vaccine BNT162b2 (Pfizer-BioNTech) according to the standard protocol (two doses of 30 µg administered 3 weeks apart). As part of the vaccination program, the vaccinated recipients were asked to report their worst local and systemic symptoms and highest body temperature that they might experience within 4 days of vaccination via a web questionnaire. To assess humoral response by the vaccine, we quantitatively measured the IgG against SARS-CoV-2 spike protein (the AdviseDx SARS-CoV-2 IgG II assay, Abbott ARCHITECT^®^) in accordance with the manufacturer’s package insert (positive threshold: ≥50.0 [AU/mL]). We also qualitatively measured the IgG against the SARS-CoV-2 nucleocapsid protein (Abbott ARCHITECT^®^; positive threshold: ≥1.40 [S/C]) to exclude those with previous undiagnosed SARS-CoV-2 infection. We measured the antibodies using blood samples donated 7, 39, and 60–74 days after the second vaccination. The study protocol was approved by the Ethics Committee of NCGM, Japan (the approved number: NCGM-A-004175). Written informed consent was obtained from all participants.

We used linear regression models to estimate the means of log_10_-transformed spike IgG titers at 7, 39, and 60–74 days after the second dose of the vaccine according to the body temperature (<37.5□, 37.5–37.9□, 38.0–38.4□, or ≥38.5□) with adjustment for age (continuous) and sex, and then back-transformed these values to present the geometric means. We also calculated the mean spike IgG titers according to the grade of local or systemic reactogenicity (Supplementary Appendix 2), with reference to the U.S. Food and Drug Administration guidance [10]. As a sensitivity analysis, we ran the above model after excluding participants with the prophylaxis use of antipyretics on vaccination (n=5).

## Results

The mean age (standard deviation) was 43 (12), and 64 % were women. The major occupations of the members were administrative staff (20 %), nurses (15 %), doctors (8 %), allied health professionals (5 %), and others (52 %). None of the participants had a history of COVID-19 and showed positive nucleocapsid/spike IgG antibody test results, removing the possibility of a previously undiagnosed SARS-CoV-2 infection.

All participants were seropositive for spike IgG from 7 through 60–74 days after the second dose of the vaccine. Participants who had a fever of 38.0–38.4□ and ≥38.5□ had significantly higher spike IgG titers at 7 days after the second dose (Figure 1) compared with that in the participants who reported no or slight fever (<37.5□) after the second dose of vaccination. The spike IgG titers tended to decrease with time among all the participants, but participants who experienced a high fever had higher titers than those who had no or slight fever even at both 39 and 60–74 days. The grades of local or systemic reactions were not significantly associated with spike IgG titers (Supplementary Table 1). These associations were virtually unchanged after excluding those with the prophylaxis use of antipyretics on vaccination.

**Figure 1.**
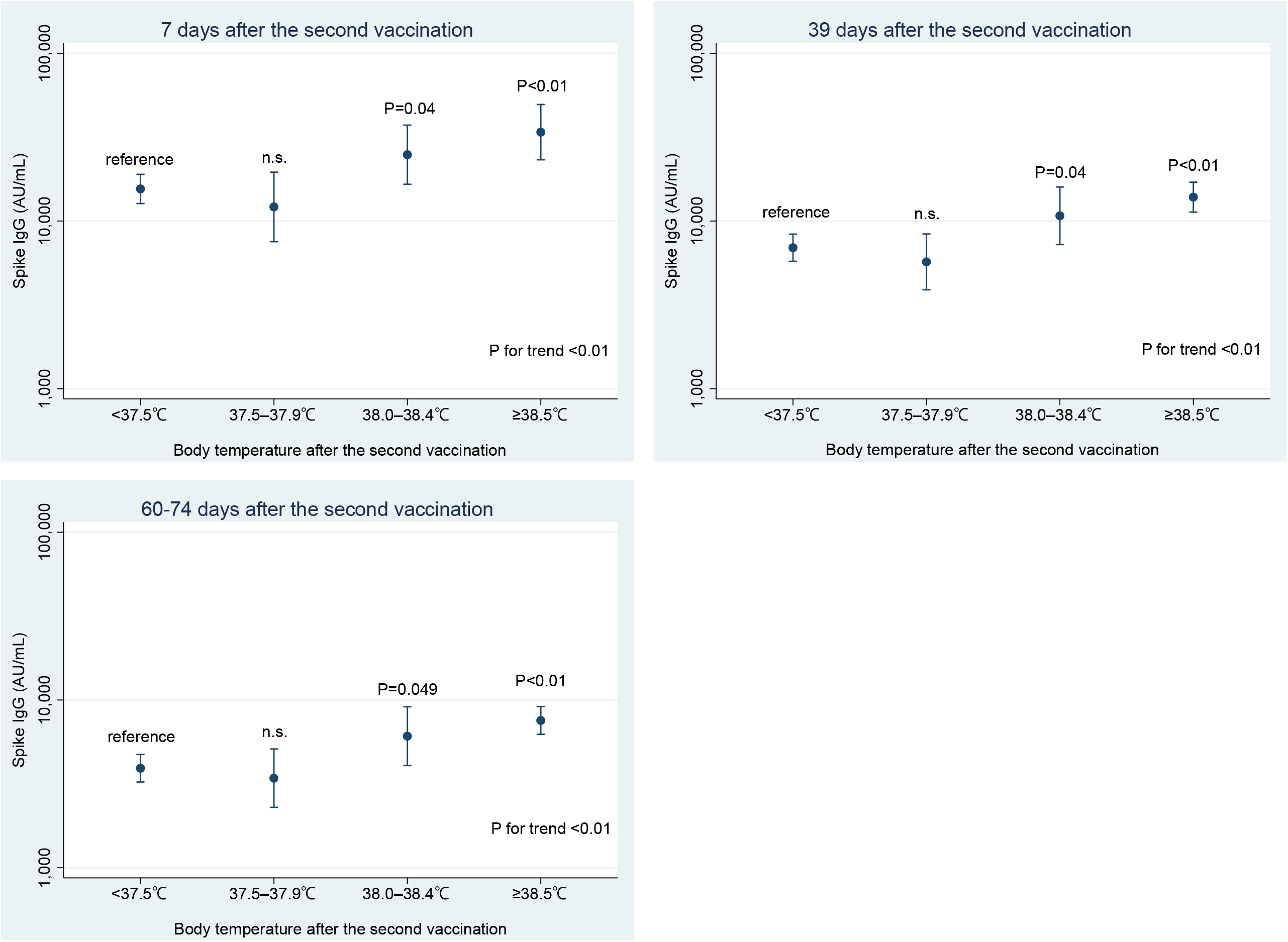
Estimated geometric means of severe acute respiratory syndrome coronavirus 2 (SARS-CoV-2) spike IgG titers with 95 % confidence intervals (CIs) by body temperature after the second dose of vaccination. Data are shown as geometric means with 95 % CIs estimated by the linear regression model adjusting for age (continuous) and sex. The number of participants in the categories of <37.5□, 37.5–37.9□, 38.0–38.4□, and ≥38.5□ were 58, 12, 12, and 6, respectively. The *P* value for trend was calculated by treating the categorical variable as a continuous term in the model. n.s.: non-significance (*P*>0.05).

## Discussion

To the best of our knowledge, this is the first study that reported higher antibody titers among recipients who experienced high fever following the two-dose BNT162b2 vaccine. This result is consistent with biological mechanisms; type-I interferon is produced as a response of innate immunity after vaccination, triggering adaptive immune response including the differentiation of T follicular helper cells, which promote B cell differentiation into antibody-secreting plasma cells [4]. Additionally, type-I interferons and other cytokines cause various side reactions, including fever. The onset of fever itself is hypothesized to augment the adaptive immune response [8]. A fever, which is more frequently observed after the second dose of vaccines [1, 2], may enhance the activation of B cells generated by the first dose of vaccine.

To date, the epidemiological data on the association between overall systemic reactions regarding the COVID-19 vaccines and antibody production are inconsistent [5-7]. In the present study, we found no association between systemic reaction gradings and the spike IgG titers. According to the clinical trial of the BNT162b2 vaccine,[1] one-fourth of the placebo recipients reported systemic symptoms (fatigue 23 % and headache 24 %) after the second dose, while only 0.8 % of the recipients reported a high fever of ≥38.0□. Thus, we speculate that symptom-based grading of vaccine side reactions may be a less sensitive variable to capture the association, if any, in the study of immunogenicity.

In conclusion, a fever of 38□ or higher after the second dose of the mRNA-based SARS-CoV-2 vaccine BNT162b2 is associated with higher concentrations of circulating spike IgG titers. Further studies are required to examine whether a post-vaccination fever can be used in the prediction of long-term immunogenicity.

## Data Availability

The datasets generated and/or analyzed during the current study are not publicly available due to ethical restrictions and participant confidentiality concerns.

## Acknowledgments

We thank the members of the working group of this study (Yusuke Oshiro, Natsumi Inamura, Haruka Osawa, and Maki Konishi) for their support.

---

**Supplementary Table 1.**
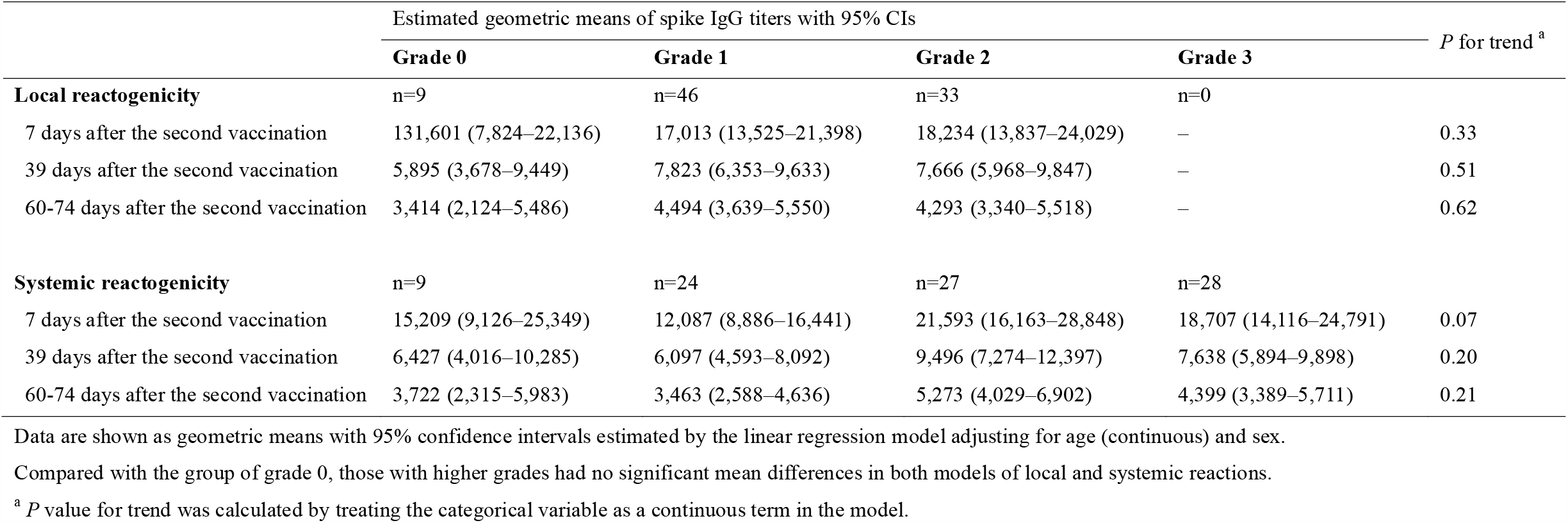
Estimated geometric means of severe acute respiratory syndrome coronavirus 2 (SARS-CoV-2) spike IgG titers with 95% confidence intervals (CIs) by the grade of local or systemic reactogenicity after the second vaccination

### Supplemental Appendix 1. Study Design

We performed a repeated survey after the start of the vaccination program in March 2021 in a cohort of the staff members of the National Center for Global Health and Medicine (NCGM), a national medical institution in Japan. We invited workers aged ≥20 years who were not taking immunosuppressive medication and who were scheduled to get the first dose of the COVID-19 vaccination on the scheduled date of the first survey (March 24 or 25, 2021). We closed the application of the study when the number of participants reached 100. Participants were vaccinated with the mRNA-based SARS-CoV-2 vaccine BNT162b2 (Pfizer-BioNTech) at NCGM according to the standard protocol (two doses of 30 µg administered 3 weeks apart). A total of five surveys were conducted for donating venous blood on the following schedules: Day 1 (immediately after the first dose), day 15 (14 days after the first dose), day 29 (7 days after the second dose), day 61 (39 days after the second dose), and day 82–96 (60–74 days after the second dose).

### Supplemental Appendix 2. The categorization of local and systemic reactogenicity

We categorized the grade of local (injection site pain, swelling, and redness) and systemic (fever, dizziness, fatigue, headache, chills, vomiting, diarrhea, muscle pain, and joint pain) reactogenicity with reference to the U.S. Food and Drug Administration guidance. Grade 0 is no adverse effect in all categories. Adverse effects were graded as follows: Without interference with daily activity (grade 1), with some interference with daily activity (grade 2), and with considerable interference with daily activity (grade 3). The score was based on specific conditions for some adverse effects: Redness (0–2.0 cm (including no adverse effects) [grade 0], 2.1–5.0 cm [grade 1], 5.1–10.0 cm [grade 2], >10.0 cm [grade 3]), swelling (0–2.0 cm (including no adverse effects) [grade 0], 2.1–5.0 cm [grade 1], 5.1–10.0 cm [grade 2], >10.0 cm [grade 3]), and fever (<38.0 [grade 0], 38.0–38.4 [grade 1], 38.5–38.9 [grade 2], ≥40 [grade 3]).

## References

1. Polack FP, Thomas SJ, Kitchin N, et al. Safety and Efficacy of the BNT162b2 mRNA Covid-19 Vaccine. N Engl J Med 2020; 383(27): 2603–15.

2. Baden LR, El Sahly HM, Essink B, et al. Efficacy and Safety of the mRNA-1273 SARS-CoV-2 Vaccine. N Engl J Med 2021; 384(5): 403–16.

3. Walsh EE, Frenck RW, Falsey AR, et al. Safety and Immunogenicity of Two RNA-Based Covid-19 Vaccine Candidates. N Engl J Med 2020; 383(25): 2439–50.

4. Teijaro JR, Farber DL. COVID-19 vaccines: modes of immune activation and future challenges. Nature Reviews Immunology 2021; 21(4): 195–7.

5. Hwang YH, Song K-H, Choi Y, et al. Can reactogenicity predict immunogenicity after COVID-19 vaccination? The Korean Journal of Internal Medicine 2021.

6. Takeuchi M, Higa Y, Esaki A, Nabeshima Y, Nakazono A. Does reactogenicity after a second injection of the BNT162b2 vaccine predict spike IgG antibody levels in healthy Japanese subjects? medRxiv (preprint) 2021.

7. Kawasuji H, Morinaga Y, Tani H, et al. Functional and quantitative evaluation of BNT162b2 SARS-CoV-2 vaccine-induced immunity. medRxiv (preprint) 2021.

8. Evans SS, Repasky EA, Fisher DT. Fever and the thermal regulation of immunity: the immune system feels the heat. Nature Reviews Immunology 2015; 15(6): 335–49.

9. Luheshi GN. Cytokines and Fever: Mechanisms and Sites of Action. Ann N Y Acad Sci 1998; 856(1 MOLECULAR MEC): 83–9.

10. Food U, Administration D. Guidance for industry: toxicity grading scale for healthy adult and adolescent volunteers enrolled in preventive vaccine clinical trials. Food and Drug Administration, US Department of Health and Human Services 2007.

